# Engineered RNA biosensors enable ultrasensitive SARS-CoV-2 detection in a simple color and luminescence assay

**DOI:** 10.1101/2021.01.08.21249426

**Authors:** Anirudh Chakravarthy, KN Anirudh, Geen George, Shyamsundar Ranganathan, Nishan Shettigar, U Suchitta, Dasaradhi Palakodeti, Akash Gulyani, Arati Ramesh

**Affiliations:** InStem - Institute for Stem Cell Science and Regenerative Medicine, Bangalore 560065; SASTRA University, Tirumalaisamudram, Thanjavur 613401; Trans-Disciplinary Health Sciences & Technology, Bangalore 560064; Red Hat, Inc., Westford, MA 01886; Department of Biochemistry, School of Life Sciences, University of Hyderabad, Hyderabad 500046; National Centre for Biological Sciences, GKVK Campus, Bellary Road, Bangalore, India 560065

## Abstract

The continued resurgence of the COVID-19 pandemic with multiple variants underlines the need for diagnostic strategies, that are easily adapatable to the changing virus. Here, we have designed and developed toehold RNA-based sensors across the SARS-CoV-2 genome for direct and ultrasensitive detection of the virus and its prominent variants. In our assay, isothermal amplification of a fragment of SARS-CoV-2 RNA coupled with activation of our biosensors leads to a conformational switch in the sensor. This leads to translation of a reporter-protein e.g. LacZ or Nano-lantern that is easily detected using color/luminescence. By optimizing RNA-amplification and biosensor-design, we have generated a highly-sensitive diagnostic assay; with sensitivity down to attomolar SARS-CoV-2 RNA. As low as 100 copies of viral RNA are detected with development of bright color that is easily visualized by the human eye, or a simple cell phone camera as well as quantified using a spectrophotometer. This makes our assay deployable all the way from a well equiped laboratory to a low-resource setting anywhere in the world. Finally, this **PHA**sed **N**ASBA-**T**ranslation **O**ptical **M**ethod (PHANTOM) using our engineered RNA biosensors efficiently detects the presence of viral RNA in human patient samples, correlating well with Ct values from RT-qPCR tests. This work presents a powerful and universally accessible strategy for detecting Covid-19 and its prominent variants. This strategy is easily adaptable to further viral evolution and brings RNA-based bioengineering to centerstage.

## INTRODUCTION

The COVID-19 pandemic has affected millions of people and caused severe disease, mortality and disruption to human activity across the world. Current estimates suggest that at least 127.8 million (as of 31 March 2021, WHO) people have been infected, and millions remain susceptible to this infection. The situation is compounded by the emergence of new variants of the virus, including potentially highly infectious strains. The COVID-19 disease is caused by a novel coronavirus SARS-CoV-2, belonging to the *Betacoronavirus* genus under the *Coronaviridae* family of viruses^1^. Due to the large numbers of potential infections, the high infectivity of the virus and the wide diversity in the clinical presentation of the SARS-CoV-2 infections, there is an ongoing need for reliable and efficient diagnostic methods. This is especially felt since a substantial portion (as much as 60-80%) of human subjects infected with SARS-CoV-2 are asymptomatic or show very mild symptoms but may still remain infectious. Further, amongst symptomatic COVID-19 patients, there is a wide variability in the nature and presentation of symptoms ^2,3^. Therefore, the direct detection of SARS-CoV-2 infections remains important. Furthermore, detection strategies need to keep up with the evolving viral variants.

Currently, diagnostic testing of human subjects for SARS-CoV-2 infections broadly rely on either RNA amplification based methods or methods for detecting the presence of viral antigens.^4–8^ The current gold standard for testing remains the reverse transcriptase-quantitative-Polymerase chain reaction (RT-qPCR) where amplification of one or more regions of viral RNA is typically detected with Taqman probes.^9–11^ Although RT-PCR based assay is considered more reliable for detection of virus, it involves significant processing steps and depends on the availability of sophisticated and expensive equipment, technical experts for instrument handling and analysis of data. Another detection method is the reverse transcriptase coupled - Loop Mediated Isothermal Amplification (RT-LAMP). In typical RT-LAMP assays, amplification of DNA from viral RNA fragments is detected using dyes sensitive to pH, DNA or pyrophosphates.^12–15^ This method is relatively faster but may generate false positives due to non-specific amplification and primer interactions. The CRISPR-Cas system has also emerged as an alternative platform for viral RNA detection. Here, CRISPR-Cas recognition of viral RNA is coupled with RNA amplification^16–19^ or used in an amplification-independent way via the use of more than one guide RNAs^20^. These are currently restricted to a lateral flow assay format for colorimetric detection or use a fluorescence read-out, which may not be lend itself to deployment in a variety of settings.

As a strategy for simple and specific SARS-CoV-2 detection that is compatible with a range of assay formats, we focused on direct detection of viral RNA fragments using an RNA biosensor approach. We searched for an RNA biosensor scaffold that was versatile, could be developed to be highly selective to SARS-CoV-2 RNA, can be used in a simple color read-out and where multiple steps of amplification built into the assay would result in high sensitivity/specificity. The previously reported Toehold RNA scaffolds met these criteria and have been widely used for detecting other viruses.^21,22^

Toehold RNAs are synthetic switches that when placed in tandem, upstream of an mRNA, can control its translation^23–28^ (Fig 1A). In a typical configuration, a toehold switch consists of a central stem containing a ribosome binding site, a translation start site and a downstream reporter gene. A part of the central stem along with a 5’ overhang is designed to specifically base pair and bind with a Trigger RNA in *trans*. In the absence of the trigger, the central, conserved stem loop sequesters the region around the RBS and start codon, not allowing translation initiation. However, binding of the Trigger RNA to the biosensor disrupts the central stem, leading to a clear conformational switch in the sensor, exposing the RBS and start codon. This leads to translation of a reporter protein such as lacZ that can be easily detected using a chromogenic substrate.

**Fig 1.**
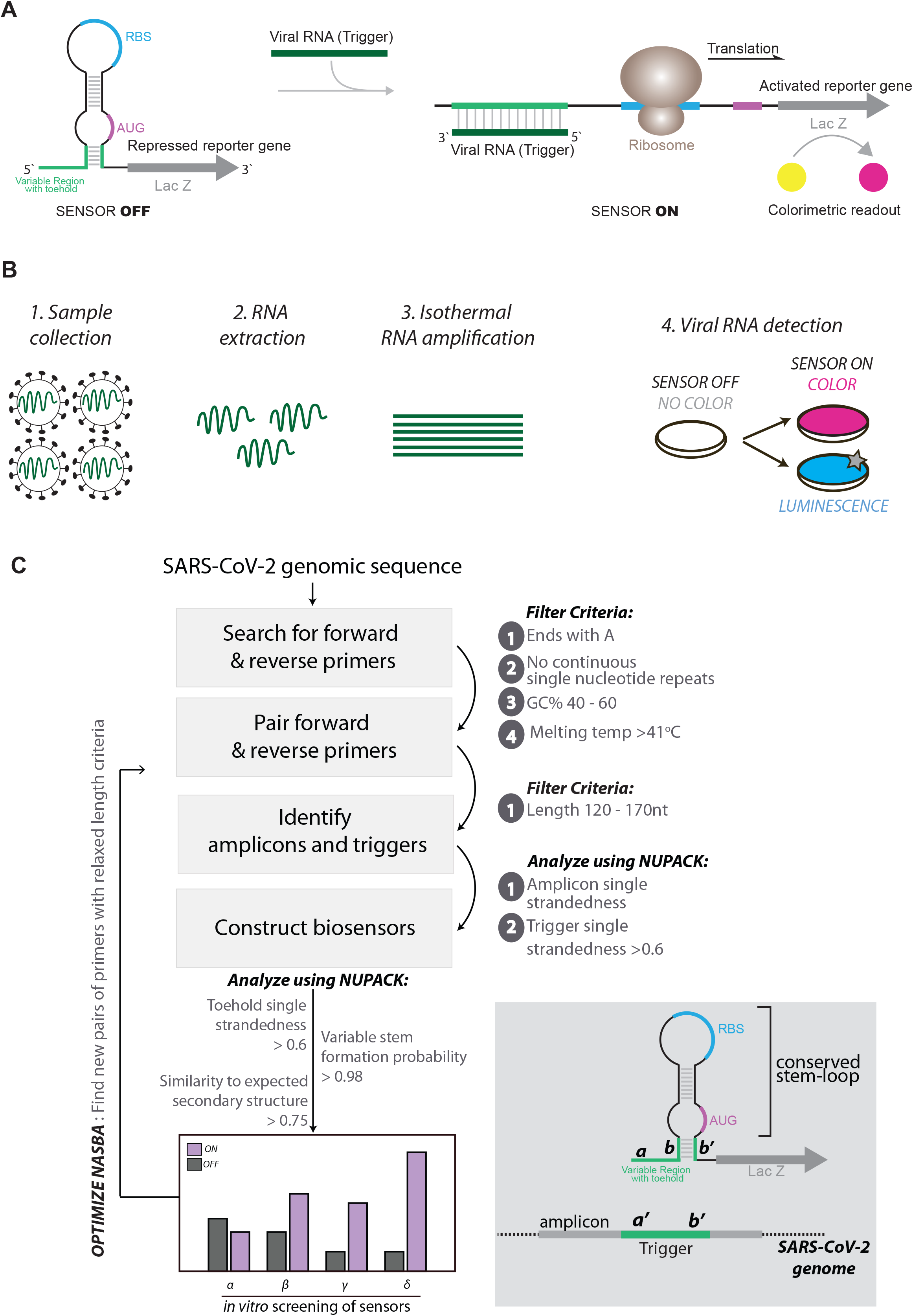
A) *Schematic of Toehold switches*. Toehold RNA switches consist of a central stem loop structure that harbors a ribosome binding site (RBS, blue) and a translation start site (AUG, pink) with a downstream reporter gene (such as *lacZ*, grey). A variable region with the toehold (green) are designed to specifically base-pair with a trigger RNA (dark green). In the absence of trigger RNA (left), the RBS and AUG are sequestered within the sensor structure and inaccessible to the ribosome. Presence of the trigger RNA (right) induces intermolecular interactions between the toehold and the trigger RNA, resulting in an alternate conformation wherein the RBS and AUG are accessible to the ribosome, enabling translation of the downstream LacZ enzyme. Production of LacZ is easily monitored with color, using a chromogenic substrate. The concept is modular and allows the use of alternate reporter genes and modes of detection. B) *Schematic showing our assay development pipeline*. RNA extracted from viral particles is amplified isothermally using NASBA (Nucleic Acid Sequence-Based Amplification) and detected with specifically designed toehold-based biosensors in an in vitro transcription-translation (IVTT) assay. The NASBA coupled IVTT assay leads to production of color that can be easily visualized by eye or with cell phone cameras or luminescence that can be quantified by luminometry. Our assay development pipeline focused on identifying targetable regions of the SARS-CoV-2 genome, design of specific biosensors, optimized primers for efficient NASBA and overall sensitivity and response of the assay. C) *Flowchart showing the bioinformatic pipeline* for primer design, and selection of biosensors. First, we searched for primers that would amplify fragments of the SARS-CoV-2 genome, with criteria as highlighted in the figure. Amplicons resulting from primer pairs were analyzed for potential Trigger regions. Amplicon and Trigger single strandedness were estimated. These Trigger regions were used to construct the biosensors which were then analyzed for toehold single strandedness, stem probability and fidelity to the expected biosensor secondary structure. Illustration (bottom right) shows the elements of the biosensor and Trigger RNA in detail.

In this study, using toehold switches as a starting point, we have engineered RNA biosensors that are highly selective for SARS-CoV-2 RNA. Isothermal amplification of SARS-CoV-2 RNA fragments, coupled with activation of our biosensors leads to production of lacZ protein. Subsequent cleavage of a chromogenic substrate results in a simple color assay for viral detection. *In vitro* characterization of these biosensors and testing of patient samples using our assay, reveals a sensitivity up to 100 copies of viral RNA, making our biosensor a feasible module for detecting SARS-CoV-2 infection in patients. Notably, our biosensor detects a region of viral RNA that is conserved across all prominent variants (such as the UK, Brazilian and South African variants). We find that this assay is compatible with different modalities of detection wherein viral RNA is detected via luminescence, in a shorter period of time. By developing new biosensors, we offer PHANTOM (**PHA**sed **N**ASBA-**T**ranslation **O**ptical **M**ethod), an ultrasensitive, highly accurate Covid detection platform that does not require any sophisticated equipment and is usable even in a low resource setting.

## RESULTS

### Analysis of the SARS-CoV-2 genome and design of potential biosensors

Toehold sensors are designed to specifically recognize and respond to a region of viral RNA (called Trigger RNA). Each biosensor consists of a sensing region contiguous with a conserved stem-loop structure that contains a ribosome binding site (RBS) and a translation start site followed by a reporter gene (Fig 1A). In the absence of viral RNA, the stem-loop sequesters the region around the RBS and the translation start site, thus keeping the biosensor “off”. Binding of viral RNA to the sensing region causes a rearrangement of the biosensor, increasing accessibility of the otherwise inaccessible ribosome binding site. This leads to increased translation of the downstream reporter gene, leading to color production.

For a toehold based biosensor to work, it requires extensive complementarity to its target viral RNA (Fig 1A). In addition to this, an ideal sensor would need to be sensitive in detection. To gain sensitivity, previous studies on toehold sensors coupled RNA amplification with sensing.^21–23^ In order to detect a wide range of SARS-CoV-2 RNA viral loads, we anticipated a need for RNA amplification (Fig 1B). Nucleic Acid Sequence Based Amplification (NASBA) is an isothermal RNA amplification method, which relies on a pair of primers and the activity of three enzymes, Reverse transcriptase, RNaseH, and T7 RNA polymerase to gain upto 10^9^-fold amplification.^29–36^

To design suitable primers for NASBA (RNA amplification), we performed *in silico* analyses of the SARS-CoV-2 RNA genome, specifically the strain MT12098.1 from India^37^ (Fig 1C). 20-to 24-nucleotide reverse (P1) and forward (P2) primers were designed to anneal throughout the viral genome. The criteria listed by Pardee et al. (2016)^21^ were adapted, wherein primers that end with ‘A’, do not have 4 or more continuous repeats of any nucleotide, have a 40-60% GC content and a Tm greater than 41°C were selected. P1 and P2 primers that anneal within 120 to 170 nucleotides of each other were paired and primer pairs were scored using the softwares Primer 3 and NUPACK. Each primer pair thus results in a 120 to 170-nucleotide region referred to as Amplicon (Fig 1C).

Within the amplicons, using sliding windows of 36 nucleotides, we searched for contiguous single-stranded regions that would be accessible to the biosensor and would serve as the Trigger RNA (Fig 1C). The complementary sequence to each Trigger RNA was then incorporated into the toehold scaffold to generate SARS-CoV-2 specific biosensors. These potential biosensors were analyzed using NUPACK, and scored on the basis of the following parameters-1) probability of formation of the lower stem (which is meant to enforce a stable “off state” of the sensor), 2) trigger single strandedness within the context of the amplicon (to increase accessibility of the trigger to bind the sensor), 3) single strandedness of the first 25 nucleotides of the sensor (meant to open the biosensor upon trigger binding) and finally the similarity to the desired consensus structure of the biosensor.

Based on these analyses we selected biosensors that 1) show high probability of forming the required sensor structure where, in the absence of viral RNA the ‘sensing region’ and its complement are base-paired and 2) the sensing region and trigger are largely open to allow for base-pairing that allows opening of the switch. 19 potential biosensors with a diversity of scores for different parameters were taken further for in vitro studies (Table S1, Table S2, Fig. 2A-B). Overall our computational pipeline yields a repertoire of biosensors capable of detecting different regions spread across the viral RNA genome.

**Fig 2.**
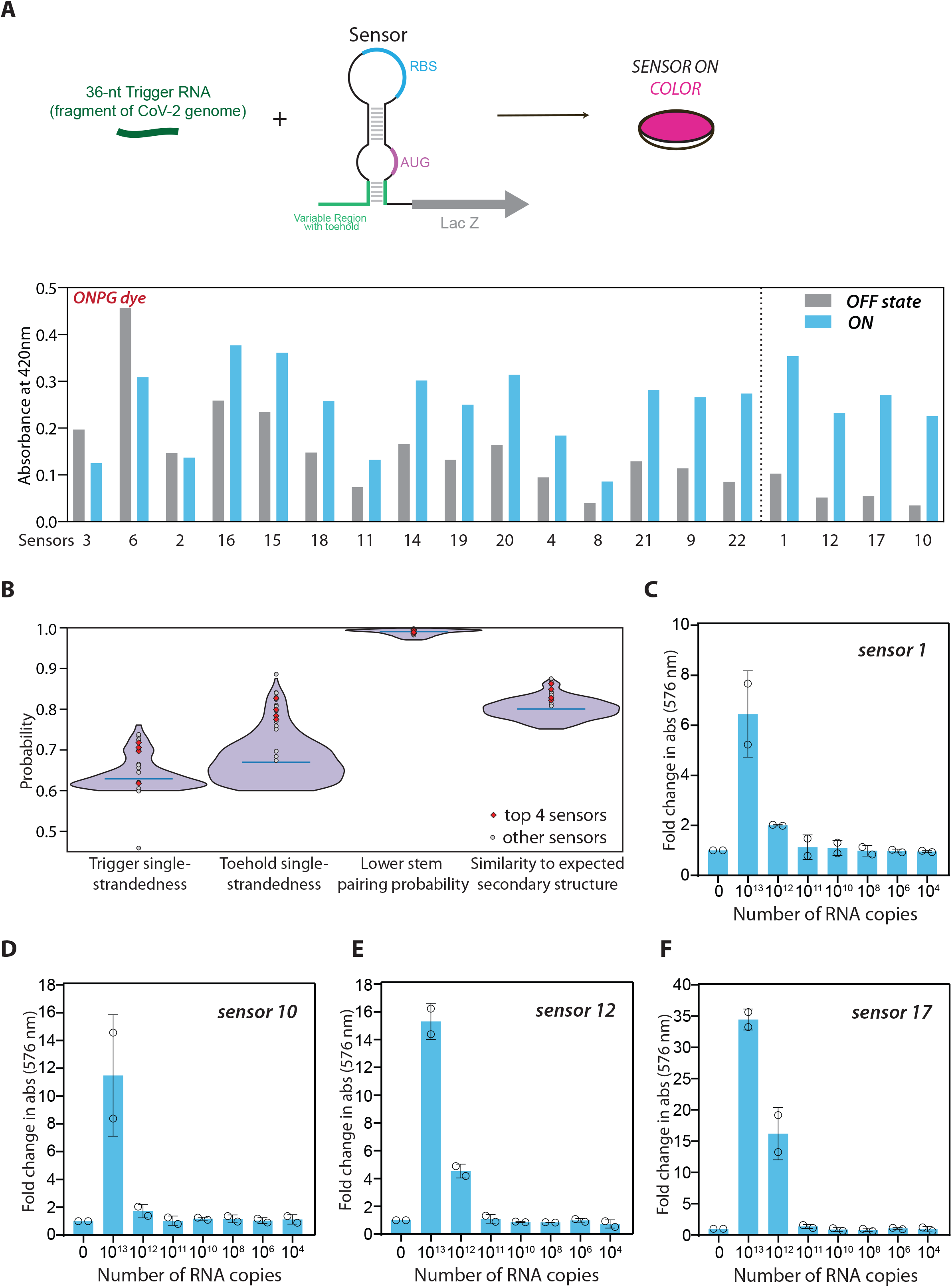
Screening and selection of SARS-CoV-2 responsive biosensors. A) IVTT assay performed on 19 shortlisted sensors was monitored using ONPG, a chromogenic substrate for lacZ. Absorbance (420nm) is plotted for each sensor, in the presence (blue) or absence (grey, OFF state) of trigger RNA. Dotted line separates sensors 1, 10, 12 and 17 which show the maximum change with respect to the off state. B) Results of the bioinformatic analysis are shown. Violin plot shows the probability distribution of different sensors with respect to 4 parameters-trigger single-strandedness, toehold single-strandedness, probability of formation of the central stem and similarity to the overall expected secondary structure of the biosensor. 19 sensors were chosen for initial screening, based on their diversity of scores (grey circles). The 4 best sensors (red diamonds) were further tested. C-F) IVTT assays performed on 4 selected sensors (1, 10, 12, 17) were monitored using CPRG, a red-shifted, chromogenic substrate for lacZ. Colorimetric response was monitored as a function of respective Trigger RNA concentration. Data shown are from two independent experiments (n=2). Fold change in absorbance (576nm) is plotted for each sensor, with varying amounts of trigger RNA (0 to 10^13^ copies of RNA). Fold change is calculated relative to OFF state of sensor (sensor alone, no RNA added). These data reveal a clear threshold RNA concentration at which the sensors are able to respond to the trigger RNA.

### Screening and identification of biosensors that detect SARS-CoV-2 derived RNAs

To test which of the designed sensors respond to their respective Trigger RNAs (synthetic RNAs identical to a portion of SARS-CoV-2 RNA), we used an *in vitro* transcription-translation (IVTT) coupled assay. In the absence of trigger RNA the sensing region of the biosensor would base-pair with its complementary sequence, keeping the RBS and translation start codon inaccessible, hence keeping the sensor in its OFF state. Presence of the Trigger RNA would sequester the sensing region, thus exposing the RBS and start codon to enable translation of the downstream lacZ mRNA. This trigger RNA-dependent production of lacZ protein is detected using colorimetry.

We tested 19 predicted sensors for their ability to produce color in the presence of Trigger RNA (Fig 2A). DNA corresponding to the biosensor was generated with a T7 promoter site at the 5’ end. This DNA when used as a template in the IVTT assay transcribes the RNA biosensor *in situ*. IVTT performed in the absence and presence of trigger RNA were compared on the basis of Absorbance at 420nm, which reports on the extent of cleavage of Ortho-Nitrophenyl-β-Galactoside (ONPG), a lacZ substrate.

We find that all of the tested sensors showed absorbance 420nm in the presence of Trigger RNA. However, 11 of these sensors also showed detectable absorbance (>0.1 A_420_) in the absence of Trigger RNA, suggesting leaky expression of *lacZ* and potentially unstable “off” state RNA conformations for this subset of sensors (Fig 2A). Notably, 8 sensors show low absorbance in the absence of trigger RNA and a significant increase in absorbance in the presence of trigger RNA. The best sensors (1, 10, 12 and 17) with the maximum fold-change in absorbance (i.e. minimal *lacZ* expression in the “off” state and a substantial increase in *lacZ* expression in the presence of trigger RNA) were chosen for further analyses. We tested if this colorimetric assay works with ChlorophenolRed-β-D-galactopyranoside (CPRG), a more sensitive, red-shifted substrate of lacZ.^38^ Our data reveals that all 4 chosen sensors 1, 10, 12 and 17 work not only with ONPG but also with CPRG, showing a greater fold change with CPRG (ranging from 6 to 35-fold increase in absorbance; Fig S1A). Hence all further experiments were performed with CPRG as the substrate.

We next examined our sensors for sensitivity of detection. Sensors 1, 10, 12 and 17 were used as template in IVTT assays in the presence of increasing amounts of trigger RNA (Fig 2C-F). These sensors show sensitivity towards 10^12^ to 10^13^ copies of trigger RNA. These experiments show that our sensors respond to their respective trigger RNAs, with a clear sensitivity threshold (Fig 2C-F).

### Isothermal amplification of RNA to enable sensitive detection by biosensors

The inherent sensitivities observed for our sensors do not lie in a range that may be useful for unaided detection of SARS-CoV-2 RNA from infected patients’ samples. For example, while viral loads are subject to much variation across populations and nature of infection,^39^ viral loads ranging from 10^8^ copies/mL to 10^3^ copies/mL (at the limit of detection for RT-qPCR based testing) have been observed in naso-pharyngeal swabs of patients.^40^ Mean viral loads observed in nasopharyngeal swab samples and saliva samples are around 10^5^ copies/mL approximately.^41^ In order to use these sensors as diagnostic tools to detect SARS-CoV-2 infection, we coupled the IVTT assay with RNA amplification. This way the RNA to be sensed is amplified to amounts that are detectable by the biosensors. Nucleic Acid Sequence Based Amplification (NASBA) is an isothermal RNA amplification method that can be coupled with toehold sensors (schematic in Fig 3A). Here, RNA (such as the viral genomic RNA) acts as template for reverse primer (P1) binding, which initiates reverse transcription at a particular position. cDNA first strand synthesis and removal of the template RNA strand by RNaseH enables binding of the forward primer (P2) which is designed to contain a T7 promoter region. This allows synthesis of the second strand of DNA. The resulting double stranded DNA product is transcribed wherein each resulting RNA (RNA amplicon) once again serves as template for P1 binding and subsequent amplification.

**Fig 3.**
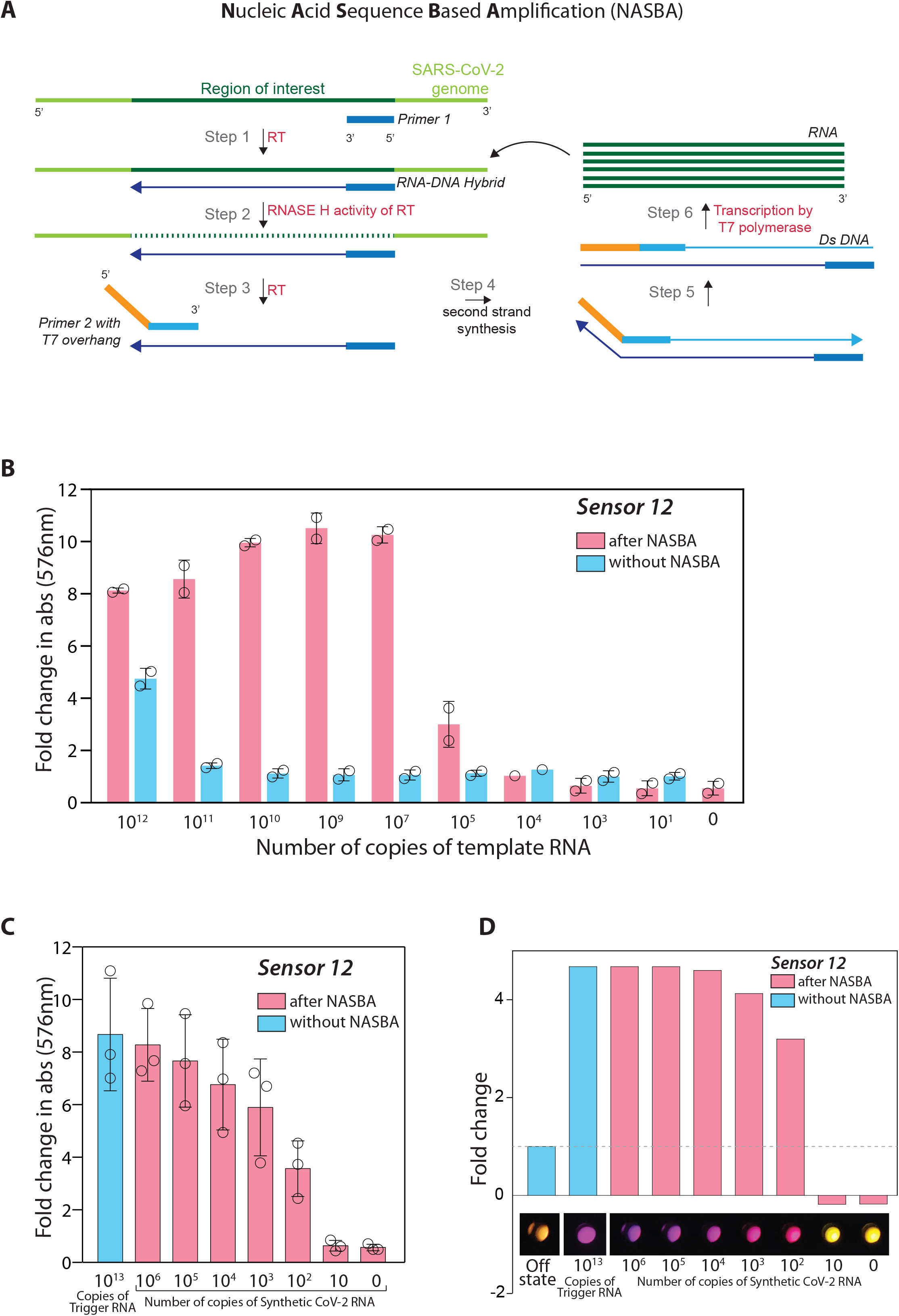
Isothermal RNA amplification (NASBA) allows for sensitive detection of SARS-CoV-2 RNA. A) Schematic for NASBA showing the various steps involved. RNA template is reverse transcribed by Primer P1 to form the first strand of cDNA. This is recognized by primer P2 containing a T7 promoter sequence. Following second strand DNA synthesis, the resultant double stranded DNA acts as a template for T7-polymerase based transcription resulting in several RNA molecules. Each newly synthesized RNA molecule in turn acts as a template for the next round of amplification, leading to iterative amplification. B) IVTT assay performed with sensor 12, with and without NASBA amplification is shown. A synthetic RNA fragment of SARS-CoV-2 containing the trigger for sensor 12 was tested for its ability to activate the sensor on its own or post-NASBA amplification. Data shown are from two independent experiments (n=2). Fold change in absorbance (576nm) is plotted against varying amounts of initial RNA (0 to 10^13^ copies of RNA), with (pink) and without (blue) NASBA. ‘0 RNA copies’ denote no template control (primer + NASBA reaction mix, no RNA added. Fold change is calculated relative to OFF state (sensor alone). C) NASBA coupled with IVTT assay performed using a commercially available synthetic SARS-CoV-2 viral RNA control (Twist Biosciences) as template. Fold change in absorbance (576nm) is plotted against increasing amounts of viral RNA (10 to 10^6^ copies of RNA). Data shown are from three independent experiments (n=3), with mean and standard deviation. Trigger RNA at 10^13^ copies (blue) is used as a positive control. Results show a sensitivity of the assay down to 100 copies of viral RNA. D) Samples from panel C were imaged using a cell phone camera. Representative image shows bright and discernable color even when starting with 100 copies of viral RNA as input. Color from individual wells was quantified and plotted as fold change relative to the off state.

In order to assess the sensitivity of our assay when coupled with NASBA amplification, we used a 136 nucleotide RNA fragment of the SARS-CoV-2 genome that encompassed the trigger for sensor 12 as a template. Increasing amounts of this RNA fragment were subjected to NASBA amplification followed by IVTT (Fig 3B). We find that when coupled with NASBA amplification, there is clearly detectable increase in absorbance even with 10^5^ copies of the RNA fragment. In stark contrast, without NASBA amplification, a minimum of 10^12^ copies of the same RNA is required to elicit a color change. Coupling with NASBA amplification thus appears to increase the sensitivity of our assay with sensor 12 by nearly 10^7^-fold, bringing these sensors into the realm of useful detection strategies for SARS-CoV-2 RNA in patient samples.

A key feature of NASBA that determines its efficiency is the selection of suitable primer pairs. We therefore further mined our list of NASBA primers in the region around the trigger for sensor 12. Here we looked for primers that would generate amplicons sized 90 to 250 while encompassing the trigger for sensor 12. These additional primer pairs were first screened for their ability to amplify a template RNA at 10^8^ copies (Fig S2A). Successful primer pairs were shortlisted and further screened for their ability to amplify 10^4^ copies of template RNA (Fig S2B). The best primer pair was used in a NASBA reaction coupled with IVTT to evaluate the overall sensitivity of the assay (Fig 3C). For this test, we moved to using the widely accepted, commercially available SARS-CoV-2 Synthetic RNA (Twist Biosciences) as template. Our results show that the best primer pair increases the efficiency of NASBA so that the effective sensitivity of our assay with sensor 12 is 100 copies of SARS-CoV-2 Synthetic RNA. A notable advantage of our assay is the facile color-based read-out that is amenable to easy detection and quantification. We find that the color produced in response to even 100 copies of RNA is easily detected with a basic cell-phone camera (Fig 3D). Put together, these data highlight that combining a suitable primer pair with our biosensor enables an ultrasensitive response to even small numbers of viral RNA copies, that is easily visualized. Bioinformatic examination of the trigger region recognized by sensor 12 and the regions recognized by the NASBA primers reveals strong conservation among all reported SARS-CoV-2 sequences, especially the prominent variants reported for SARS-CoV-2 (the UK, South African and Brazilian variants). This implies that our detection strategy would work for these key variants.

### Luminescence detection accelerates assay response to SARS-CoV-2 RNA

The biosensor design used here is modular and amenable to diverse read-outs, wherein the reporter gene can be switched from one to another (Fig 4A). The lacZ based readout used thus far allows for easy visualization of color in a sensitive manner. An important aspect of a diagnostic assay is the time taken to build a measurable response. To address this, we used the SARS-CoV-2 Synthetic RNA (Twist Biosciences) as input for NASBA and monitored the kinetics of the IVTT reaction (post-NASBA). We see a clear graded response to the copy number of RNA, with a faster build-up of color for higher initial RNA concentrations. 10^6^ copies of RNA show discernable color (A_576_>1) even at ∼60 minutes while 100 copies of RNA are detectable at 100 minutes (Fig 4B). We tested the response of a luminescence based biosensor by replacing lacZ with the Nano-lantern protein, a fusion of Renilla Luciferase8 and the mTurquoise2 fluorescent protein (Fig 4A,C). We find the sensitivity of our assay remains conserved with detection of 100 copies of initial RNA template. Notably, the response is accelerated and even in 30 minutes we are able to detect substantial build up of luminescence with 100 copies of RNA (Fig 4C). These results confirm that our SARS-CoV-2 biosensor is compatible with diverse read-outs, which can be utilized based on equipment availability and local needs.

**Fig 4.**
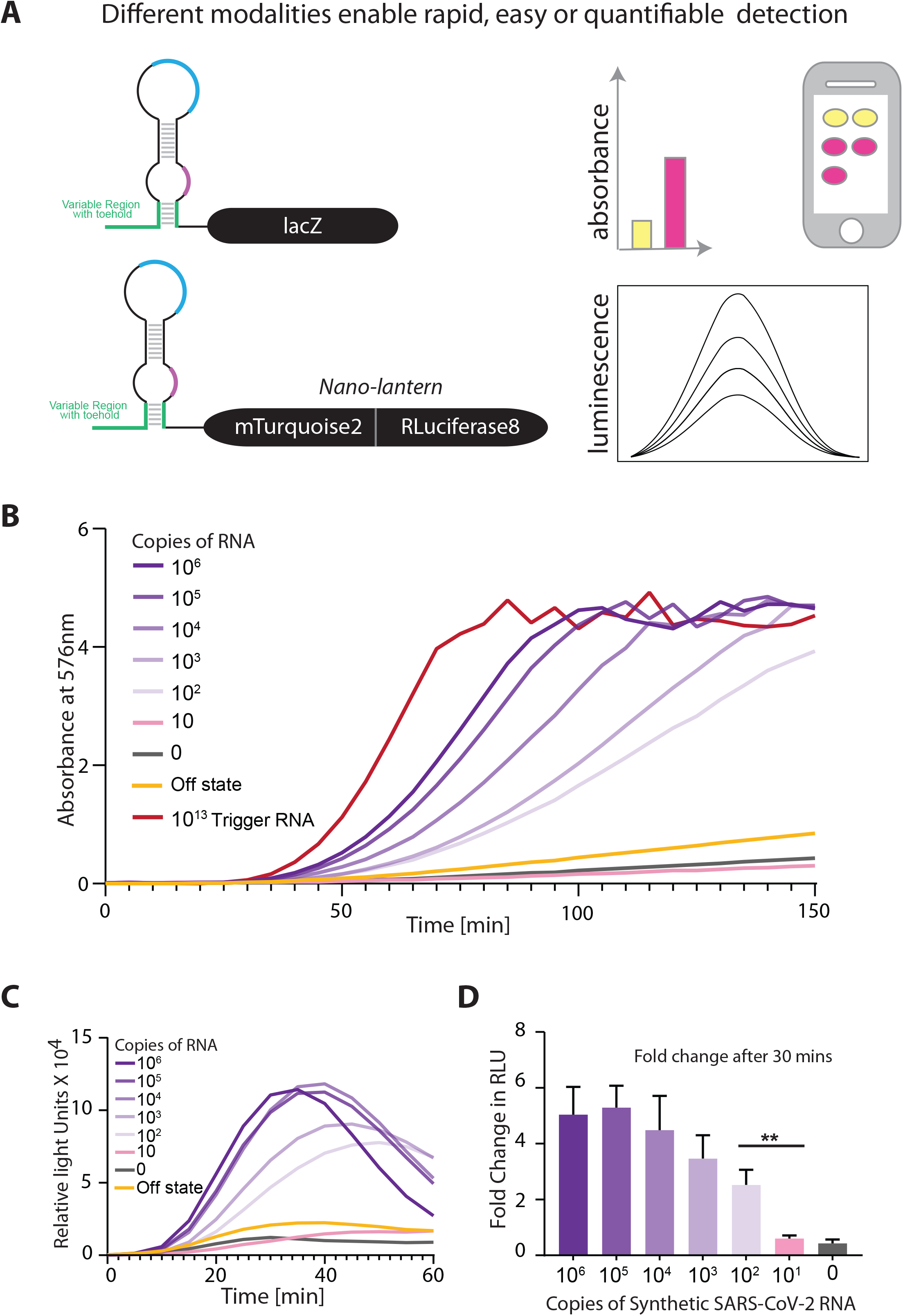
Luminescence read-out speeds up detection of SARS-CoV-2 RNA. A) Illustration shows the construction of biosensors with alternate modalities of viral RNA detection. Biosensors may comprise of a color-producing enzyme like lacZ or a luminescence producing system coupled to the RNA detection sensing module. For luminescence based detection, the Nano-lantern system (fusion of Renilla luciferase8 with mTurquoise2 fluorescent protein) was used. B-C) NASBA coupled with IVTT assay performed using a commercially available synthetic SARS-CoV-2 viral RNA control (Twist Biosciences) as template. Sensor 12 fused to lacZ was used for IVTT. Representative data from 3 independent experiments (n=3) are shown. Panel B shows Absorbance (576nm) over time for varying amounts of viral RNA (0 to 10^6^ copies of RNA). Panel C shows relative luminescence intensity over time for varying amounts of viral RNA (0 to 10^6^ copies of RNA). Trigger RNA at 10^13^ copies (red) is used as a positive control. For luminescence readout, sensor 12 was fused to the Nano-lantern reporter. D) Fold change in luminescence intensity for varying amounts of viral RNA (0 to 10^6^ copies of RNA) after 30 minutes of the IVTT reaction post-NASBA. Data shown are from 3 independent experiments. Statistical significance (p-value <0.01 is shown as ‘**’ and was calculated using two-tailed unpaired t-test.

### Detecting SARS-CoV-2 RNA in patient samples

Having established assay sensitivity down to 100 RNA copies for sensor 12, we checked if this biosensor could detect SARS-CoV-2 RNA in patient samples (Fig 5A, Fig S3). To this end we sampled RNAs extracted from nasopharyngeal swabs of 47 human subjects, whose samples had been tested with the standard RT-qPCR method (at the inStem-NCBS Covid testing Center, Bangalore). Samples spanning a range of Ct values from the RT-qPCR test were tested with our assay. Samples with Ct values from 35 to 16 through a standard RT-qPCR assay (patients designated positive for Covid infection) showed discernable build up of color (absorbance at 576nm) in our assay (Fig 5A, Fig S3). Importantly, the color produced in these samples is bright and easily detected by eye and is quantifiable through a cell-phone camera (Fig 5B). Absorbance changes observed in our assay correlate well with Ct values from RT-qPCR, and hence correlate with the amount of viral load in the patient samples. Three samples with Ct values 36-38 were indistinguishable from samples designated negative (Ct > 40) as well as the OFF state of the biosensor, indicating the possible limit of the assay in the absorbance mode. Importantly, the 27 samples with Ct > 40 (from subjects designated negative for Covid infection) showed no significant absorbance in our assay, and no discernable color both by eye or through a cell-phone camera photo.

**Fig 5.**
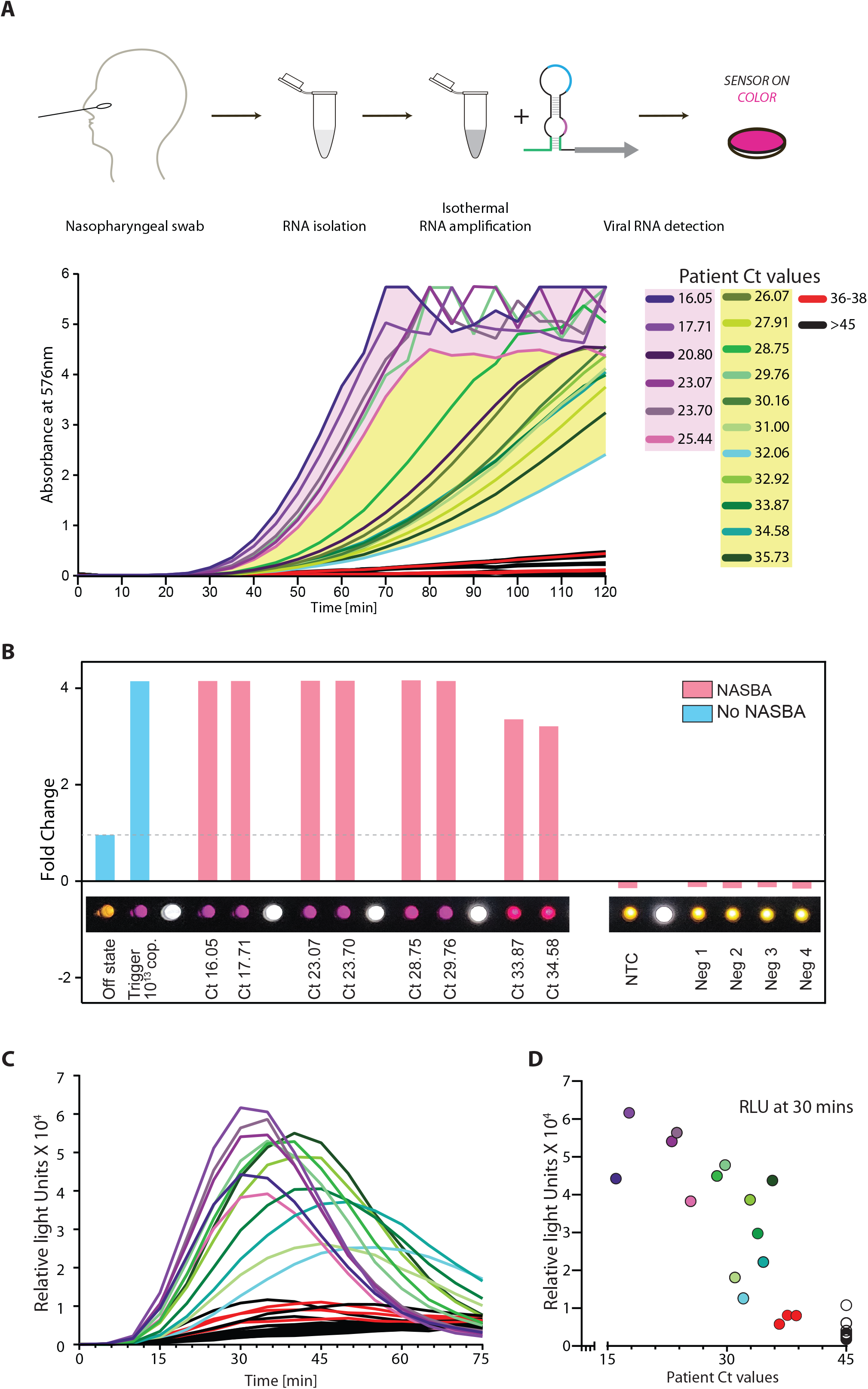
Sensitive and specific detection of SARS-CoV-2 RNA in human patient samples. A) NASBA coupled with IVTT assay performed on RNA extracted from nasopharyngeal swab samples. Samples had already been tested for the presence of SARS-CoV-2 RNA using standard real-time RT-PCR test and designated negative or positive (with accompanying Ct values shown here). Sensor 12 fused to lacZ was used for IVTT. Shown are Absorbance (576 nm) values over time. Samples are color coded based on their designation and Ct values; Ct values 16 to 25.4 (purple), Ct values 26 to 35.7 (green), Ct values 36-38 (red), -ve patients with Ct values > 45 (black). Caption indicates Ct values corresponding to individual curves. B) Photo from a basic cell phone camera showing color readouts of IVTT assay from patient samples. Assay (as described in A) performed in plate format. Bar graph shows quantitation of individual wells showing color readouts from selected patient samples. A cell phone camera image can be used to distinguish COVID-19 positive from negative samples. C) Luminescence read-out for patient samples. Sensor 12 fused Nano-lantern is used for IVTT assay. Shown are luminescence intensities for patient samples over time. Color coding of samples same as above. D) Luminescence intensity at 30 minutes of the IVTT assay (post-NASBA amplification) versus Ct-value for patient samples. Ct values were determined through the standard real-time RT-PCR test. Assay read-out clearly discriminates samples positive for viral RNA from negative samples.

In the luminescence mode, we see a similar detection of positive patient samples ranging from Ct 16 upto ∼Ct 35 and a clear discrimination from the negative samples (Fig 5C-D, Fig S4). In this range of Ct values, the signal is detectable at 30 minutes post-amplification (Fig 5C-D). For the range of Ct values 30 to 35, the signal is further enhanced between 30 to 50 minutes post-amplification (Fig 5C, Fig S4). Thus, our biosensor appears to be sensitive to the amount of SARS-CoV-2 RNA typically encountered in the population, and the readout is specific (no color/luminescence from negative samples). Based on these collective data, we propose PHANTOM (**PHA**sed **N**ASBA-**T**ranslation **O**ptical **M**ethod) as a feasible module for unambiguous Covid detection, that is universally accessible in a variety of settings.

## DISCUSSION

In this report, using computational methods, we have designed toehold RNA based biosensors that are tuned to sense different fragments of the SARS-CoV-2 RNA, spread across the genome. Extensive *in vitro* screening and characterization led us to identify biosensors that turn on translation of the reporter lacZ, in response to SARS-CoV-2 RNA fragments. Taking one of these biosensors forward, we coupled isothermal NASBA based RNA amplification to the in vitro transcription and translation assay to achieve sensitivities in the range of 100 copies of viral RNA. Alternate luminescence based detection enabled a faster time response post-amplification, highlighting the modularity of our system. When applied to patient samples, our assay provides a clear response that discriminates viral-positive from negative samples. Collectively, we present here a diagnostic platform with a read-out that is quantifiable and correlates excellently with the gold standard RT-qPCR assays. Further, this assay can be deployed in a low resource setting as the read-out is easily visualized by eye as well as through a simple cell phone camera, and the assay itself does not require any expensive equipment.

A key advance of this work is to exploit the toehold concept for SARS-CoV-2 detection. The toehold switch concept itself is an elegant strategy for RNA detection that has been used successfully for viral infections and other pathologies. A hallmark of this concept is an RNA-based switch that is designed for *specific* and *direct* detection of any RNA sequence. A key challenge in this toehold design is to ensure that the sensor is truly “off” in the absence of target RNA and turns on *only* in response to the target. We found that both the computational analyses and *in vitro* screening were crucial to overcome this challenge. We initially chose sensors with diverse scores across bioinformatic parameters. Combining this with targeted *in vitro* screening, we were able to identify the sensors that show a suitable ON to OFF state response.

The toehold switch concept is highly modular, allowing different reporters and hence diverse read-outs for detection.^21–26^ We exploited this to develop two independent modes of detection, i.e. color (using lacZ) versus luminescence (using nano-lantern). The production of bright color even at 100 copies of viral RNA allows for very easy visualization by eye, enabling a yes-no answer for the presence of viral RNA. To remove the subjective bias inherent in eye-based detection, we also show that the color produced in response to viral RNA can be recorded and validated through a basic cell phone camera. This feature of the PHANTOM assay would be extremely valuable in a low resource setting since neither conducting the assay nor interpretation of its results require specialized exquipment or training. This is aided by the fact that this assay is compatible with most formats and can be conducted in tubes, paper strips or high-throughput multi-well plates. In a laboratory setting, both the luminescence as well as colorimetric read-outs can be measured quantitatively using a luminometer or spectrophotometer. Comparing the two modes of detection, we observe a significant decrease in response time where in luminescence build up is seen even in 30 minutes post-amplification. Here, it is possible that different extent/kinetics of translation of different reporters as well as their differences in their enzymatic activities play a role in determining the assay time.

Another significant step of optimization in our assay is during RNA amplification using NASBA. Previous reports have also highlighted the importance of primer design and optimization to achieve efficient NASBA.^22,42,43^ We observed that primers with fairly similar basic criteria showed significantly different NASBA efficiencies. Therefore, after an initial round of screening for sensors, and zeroing in on an efficient sensor, we again screened for the most efficient NASBA primers for the selected sensor. This was crucial in achieving the overall sensitivity of the assay. We could detect 100 copies of viral RNA and overall sensitivity of 80 attomolar viral RNA. Our results reveal the importance of empirical screening with a diverse set of primers for efficient NASBA.

With a robust and sensitive assay in hand, we asked if the time taken for this assay could be optimized further. One of the key steps here is the NASBA amplification hence we tested to see if the time taken for NASBA could be decreased. We found that even at the limit of detection (i.e. 100 copies of viral RNA), we were able to complete the NASBA amplification step in 40 to 60 minutes (Fig S5). This, along with the faster luminescence detection effectively reduces the assay time to ∼90 minutes or better.

Finally, our results with the human patients show that our assay can clearly distinguish viral-positive from negative samples. Indeed there is strong correlation between the assay response and Ct values obtained from the RT-qPCR test. The overall sensitivity in the attomolar range ensures detection of infection in the majority of Covid-positive patients in a population. The GISAID database reports ∼842372 high quality sequenced genomes (as of 31 Mar 2021) from different clades of the SARS-CoV-2 virus. The viral RNA fragment that turns on our sensor is located in the ORF1ab (Nsp13) region of the genome. This region is completely conserved in greater than 99.1% of deposited genomes of SARS-CoV-2. This indicates that our sensors would be capable of detecting nearly all of the currently sequenced strains of SARS-CoV-2. Most significantly, this region and the regions targeted by the NASBA primers are invariant in 99.2% of the genome sequences from the newly reported UK variant and 98.5% and 99% in the Brazialian and the South African variants of SARS-CoV-2. A feature of toehold based biosensors is the multiple check-points for specificity in detection. One level of specificity comes from primers that amplify only a given region of the viral RNA and second comes from sequence-specific interactions between the viral RNA fragment and the biosensor. This is exemplified by our results wherein we observe very low false-positive rates. Detectable color is produced in positive patients (20 positive) but no color in negative patients (27 negative patients). Finally, our assay works well with the standard mode of nasopharyngeal sample collection. Combining this method of detection with other modes of sample collection such as saliva, would be a significant advance going forward.

In conclusion, our engineered biosensors along with the PHANTOM platform provide a powerful strategy for Covid detection. This can not only mitigate uncertainties in global supply chains and counter a shortage of reagents but also serve different local conditions and contexts that may benefit from diverse testing strategies.

## METHODS

### Bioinformatic analysis and sensor design

To establish a pipeline for designing SARS-CoV-2 specific toehold sensors, we started by searching for primers that would anneal to the SARS-CoV-2 genomic RNA. An Indian strain of SARS-CoV-2 (Accession Code MT012098.1) was downloaded from NCBI and used for analysis.

#### Step1. Searching for primers

Using a custom program we divided the genomic sequence (and its reverse complement) into all possible fragments of 20 to 24 nucleotides. Fragments which end with an Adenosine, do not have a continuous stretch of 4 or more of the same nucleotide, show a GC content between 40 to 60% and have a melting temperature above 41°C were shortlisted and considered as primers. Fragments arising from the sense strand were considered as forward primers (P2 primer) and were prefixed with a T7 promoter sequence(AATTCTAATACGACTCACTATAGGGAGAAGG). Fragments from the reverse complement were considered as reverse primers (P1 primer). All primers were scored using Primer3 (v. 2.5.0^44^), and NUPACK (v. 3.2.2^45^). Reaction conditions were defined as 41°C and buffer containing 50mM sodium and 12mM magnesium and primers were then scored based on the following parameters:

1. First 6-nt GC count (including hybridised region for forward primers
2. Last 6-nt AT count
3. Total %GC (including hybridised region for forward primers)
4. Single strandedness
5. Concentration of primer remaining in monomeric form
6. Single strandedness of the last 6 nucleotides
7. Contiguity length

#### Step 2: Identifying targetable regions (Trigger RNAs) within the SARS-CoV-2 genome

Forward and reverse primers that are separated by 120 to 170 nucleotides (inclusive of primer length) are paired. The region amplified by each primer pair is considered as an amplicon. Single-strandedness of all resulting amplicons was estimated using the *complexdefect* function in the NUPACK package^45^). For this amplicon analysis, we included the standard 9 nucleotides (GGGAGAAGG) appended to each sequence at the 5’end. Separately, for sensor 12 NASBA primer screening experiments (Fig S2), forward and reverse primers separated by 90 to 250 nucleotides (inclusive of primer length) were paired.

Each amplicon from the previous step was split into continuous windows of 36 nts each. Each 36nt sequence is considered a Trigger RNA. Using the *pairs* function in NUPACK, we calculated the pair probabilities for the whole amplicon. Using a custom code, we then extracted probabilities related to the Trigger regions. This Trigger single-strandedness (in the context of the whole amplicon) was considered for our analysis (Fig S2).

#### Step 3: Construction and analysis of Biosensors

To construct the biosensors, the reverse complement of each Trigger sequence was appended to the 5’ end of the conserved stem-loop in the toehold design. The sequence of the conserved stem-loop was taken from Series B toehold sensors as described in Pardee et al. 2016^21^ Thus, each complete biosensor (full sequences are given in Table S1) consists of: 5’-reverse complement of **trigger** + conserved stem-loop + first 11-nt of **trigger** + (N_1_) + linker + reporter gene (lacZ/nano-lantern),

- where (N) is any nucleotide.
- to aid in efficient transcription, we ensured that for each sensor, the T7 promoter sequence was followed by 3Gs.
- only sensors that do not possess a stop codon were considered further.

Using NUPACK, the sensors were analyzed for single strandedness of the toehold region (initial 25nts + G’s added) and the probability of formation of the lower (variable) stem (b-b* in Fig 1C). In addition, the region from the 5’end to (inclusive of) the linker was separately analyzed for its similarity to the expected secondary structure (as described in ^21^).

#### Step4: Choice of sensors for in vitro screening

Four parameters were considered while choosing sensors for *in vitro* screening. Trigger single-strandedness (>0.6), toehold single-strandedness (>0.6), probability of formation of the variable stem (>0.98), and the similarity to expected secondary structure (>0.75) were used as minimum criteria. From the list of sensors that passed this criteria, 25 sensors were chosen. Their trigger sequences were checked for potential similarity to the human genome (Accession code: GCF_000001405.38) and transcriptome (Refseq, 16May2020) using the megablast module of NCBI-BLAST, with default parameters (e-value threshold: 0.05, gap costs: creation -5 extension -2, match/mismatch score: +2/-3). Only the triggers that did not have any hits were chosen for *in vitro* analysis (sensor parameters in Table S1). 19 sensors meeting this criteria were picked for screening.

### In vitro Preparation of Toehold biosensors

The complete DNA template for an RNA biosensor consists of the T7 promoter, sensor sequence with RBS and start codon, linker and the lacZ (or Nano-lantern) reporter gene. To construct this, we first cloned the linker (this is common to all sensors) in frame with the *lacZ* or nano-lantern gene in a standard *E. coli* plasmid. Then, we purchased DNA oligos (listed in Table S4) containing the T7 promoter, sensor, RBS, the start codon and the linker. A PCR was carried out to stitch this DNA oligo (T7 promoter to linker) with the linker-lacZ DNA, using relevant primers (Table S4). This results in a linear DNA template that was purified (Promega, Cat# A9282) and subsequently used as input to the IVTT assays.

### In vitro transcription of Trigger RNAs and template RNAs

The RNA templates for NASBA reactions and relevant cell free *in vitro* transcription and translation reactions were synthesised by *in vitro* transcription reactions. This was done in a 40µL in vitro transcription reaction system. Each reaction contained 2.5µg of the relevant DNA template, 4µL of 10X T7 polymerase reaction buffer (Toyobo, Cat#TRL-201), 5.5µL of 50mM MgCl_2_, 4µL of 25mM rNTPs (NEB, Cat#N0450S), 2µL (100 units) of T7 RNA polymerase enzyme (Toyobo, Cat#TRL-201), 2 µL (0.2 units) Yeast Inorganic Pyrophosphatase (NEB, Cat#M2403S), 0.5µL (20 units) RNaseOUT (Invitrogen,Cat#10777019) and the remainder of the reaction volume was made up to 40µL with nuclease free water and incubated at 37°C for 2 hours. Following this, samples were treated with 2µL (4 units) of DNAse I enzyme (NEB, Cat#M0303S) at 37°C for 1 hour and purified using the ZymoResearch RNA Clean and Concentrator RNA purification kit (Cat# R1015). Trigger RNA products were not purified using the kit but were run on a 6% urea polyacrylamide gel and visualised by UV shadowing. The appropriately sized bands were excised out of the gel and the RNA was recovered by passive elution. The RNA was then precipitated with ethanol, centrifuged and the pellet re-dissolved in nuclease free water for further use.

### In vitro transcription translation assay

Cell free *in vitro* transcription and translation reactions (NEB PURExpress, Cat#E6800L) were prepared using the manufacturer’s protocol. In a total reaction volume of 10µL, 4µL of solution A was added along with 3µL of solution B, 0.25µL (10Units) of RNase Inhibitor (Thermofisher-scientific,Cat#10777019) and 125ng linear DNA template. Reactions were initiated with trigger RNA or NASBA product wherever applicable and incubated at 37°C for 2 hours. For IVTT reactions with trigger RNA (Fig 2A), 1µL of each 9µM Trigger RNA stock was added to 10µL cell free reaction with respective sensors. For trigger RNA dilutions (Fig 2C-F), a stock of trigger RNA was prepared and serially diluted to obtain concentrations from 9µM to 9fM. These Trigger RNA concentrations were added to approximately obtain copies of 10^13^ - 10^4^ in each reaction tube.

For reactions with Ortho-Nitrophenyl-β-Galactoside (ONPG)(Sigma, Cat#N1127) substrate, after incubation at 37°C for 2 hours, 0.5µL of 10mg/mL ONPG dye was added to the reaction mix and incubation continued at 30°C for 15 minutes. The reaction was quenched with 2µL of 2M Na_2_CO_3_. Absorbance (420nm) was recorded for the samples (at a dilution of 1:10), using a cuvette of pathlength 1mm, on a Spectrophotometer (Eppendorf Biospectrometer). Fold change in absorbance was calculated relative to the sensor “OFF” state.

For reactions with ChlorophenolRed-β-D-galactopyranoside-CPRG (Sigma, Cat#59767), 0.75µL of 12mg/mL substrate was added from the start of the IVTT reaction and incubated at 37°C for 2 hours. Samples were quenched with 2µL of 2M Na_2_CO_3_ absorbance recorded at 576nm.

Alternately, we performed IVTT reactions in 384-well plates (Corning, Cat# 3544). Here, total IVTT reaction volume was proportionately reduced to 5µL. Reactions were set up as described above and the plates were placed in a Varioskan Lux instrument (ThermoScientific) set at 37°C. Absorbance was monitored at 576nm, at 5 minute intervals, for 150 minutes. The linear measurement range of the

Varioskan Lux multiplate reader is 0-3 absorbance for a 384 well plate. The number of replicates for each experiment is indicated in the individual figure legends. Information on statistical tests carried out and statistical measures plotted are also indicated in the individual figure legends. All absorbance based plate reader experiments were first baseline corrected using a blank sample and each sample was normalized such that the lowest absorbance measurement was set to 0. All data was plotted using GraphPad Prism 8 and figures were made using Adobe Illustrator.

### Mobile phone image acquisition and analysis

Post IVTT reaction, the Corning® 384 well clear bottom microplate was placed upside down on a white LED light source. An RGB image was acquired using a smart phone camera (Xiaomi PocoF1, Redmi Note 9 Pro Max). Image was further processed and analysed using Fiji/image J 1.52p.^46^ First, the RGB image was cropped into the required dimension. Second, the cropped RGB image was split into three independent 8-bit greyscale images (Red, Green, Blue components of the original image). Third, a uniform circular region of interest (ROI) was drawn within the area of each well (green channel grayscale image), to determine the signal by averaging the pixel values. Blank well value was subtracted from all other well values. Finally, the fold change was quantified by using the average signal from sensor offstate well.

### Luminescence assays

IVTT reactions were set up in 384-well plates as described above and monitored using the Varioskan Lux instrument (ThermoScientific) set at 37°C. Intensity was measured in the ‘normal’ luminescence mode without wavelength selection.

### Isothermal RNA Amplification (NASBA)

For NASBA reactions, a relevant RNA template (sequence in Table S4) from SARS-CoV-2 genome was made using in vitro transcription as described above. In a reaction volume of 17.7µL, RNA template (ranging from 10 to 10^13^ copies) was incubated with 4µL of 5X AMV RT Buffer (Roche, Cat#10109118001), 1.6µL of 50mM MgCl_2_, 2µL of 25mM rNTPs (NEB, Cat#N0450S), 2µL of 10mM dNTPs (NEB, Cat#N0447S), 0.18µL of 1M DTT (VWR,Cat#3483-12-3) 3µL of 100% DMSO (Sigma, Cat#D8418-50ML), 0.2µL of 10mg/mL BSA (Roche,Cat#10735078001), 0.5µL each of 10µM forward and reverse primers. The assembled reaction mix was initially incubated at 65°C for 5 minutes, followed by 50°C for 5 minutes, post which an enzyme mix containing 1µL (50 Units) T7 RNA Polymerase (Toyobo, Cat#TRL-201), 1µL (20 Units) AMV-RT (Roche, Cat#10109118001) 0.1µL (0.2 Units) RNaseH (Roche, Cat#10786357001) and 0.2µL (12.5 Units) of RNaseOUT (Invitrogen, Cat#10777019) was added and the reaction was incubated at 42°C for 2 hours. Post-NASBA, the samples were either stored at -80°C or taken further for IVTT assays.

For the initial NASBA experiments (Fig 3B), an RNA template of 127 nucleotides (corresponding to coordinates 17673 - 17799 of MT012098.1 accession code) was used at different starting copy numbers (10 to 10^12^). A sequence of GGGAGAAGG was appended to the 5’ to increase transcription efficiency. For the NASBA primer screening experiments (in Fig S2), an RNA template of 3.1kb (corresponding to coordinates 17131 - 20234 of MT012098.1 accession code) was used initially at 10^8^ and subsequently at 10^4^ copies. For testing the sensitivity of the best NASBA primer pair a commercially available synthetic SARS-CoV-2 RNA (Twist Bioscience, Cat# 102024) was used. Primers used in each of these experiments are listed in Table S3). Design and rationale behind the choice of primers is described in the Bioinformatics section above.

A total of 28 primer sets were selected for sensor 12. Of these, 14 primer sets contained a forward primer (P2) with a minimal T7 promoter (TAATACGACTCACTATAGG), and are referred to as S01 to S14 primer sets. The remaining 14 primer sets contained a longer T7 promoter with a purine stretch (AATTCTAATACGACTCACTATAGGGAGAAGG) as used in Dieman et al (2002)^47^, and are referred to as L01 to L14 primer sets. The primer screen was carried out in two phases. The first phase involved the screening of all primer sets with 10^8^ copies of RNA as starting material. This was done to shortlist all primer sets that were capable of amplifying the target RNA to enable their detection using our IVTT assay. From these experiments, we shortlisted the primer sets that showed the fastest development of colour in our IVTT assays. A total of 11 primer pairs were shortlisted for the second phase of our screen. These primers were tested with only 10^4^ copies of template RNA in order to identify primer sets that could better the sensitivity of our previously used primers. The primer set (Primer pair: S01) that showed the fastest development of colour and the highest signal to noise ratio was finally selected.

## Data Availability

The datasets generated during and/or analyzed during the current study are available from the corresponding authors upon reasonable request

## ACKNOWLEDGEMENTS

We are grateful to Professor Satyajit Mayor for unstinting support for this project and very insightful discussions. We also thank Prof. Mayor for help with continued funding for this project. We especially thank the inStem-NCBS testing facility for patient samples. We thank Prof. Apurva Sarin for support. We are grateful to the Azim Premji Philanthropic initiative (APPI), PNB Housing and IQVIA for funding. We acknowledge support from the Department of Atomic Energy, Government of India and the National Centre for Biological Sciences-TIFR, under project no. 12-R&D-TFR-5.04-0800. Research in the A.R. lab is also supported by the Human Frontier Science Program research grant RGY0077/2019 and DBT/Wellcome Trust-India Alliance (IA/I/14/2/501521). A.G. thanks University of Hyderabad, the School of Life Sciences and Department of Biochemistry (UoH) for support. We are grateful to Professor Dayananda Siddavattam for support and discussions. A.G. also thanks Professor Krishnaveni Mishra for support. Research in AG lab is also supported by Scientific and Engineering Research Board (SERB), DST, Govt. of India (EMR/2017/005093).

## ETHICS STATEMENT

All clinical patient samples from nasopharyngeal swabs, which underwent real time RT-PCR testing according the ICMR guidelines, were provided by the inStem-NCBS COVID testing center. Approvals by the Institutional Human Ethics Committee (NCBS/IEC-17/002; inStem/IEC-17/001), the Institutional Biosafety Committee (TFR:NCBS:31-IBSC_UR; inStem/G-141(3)-15/CJ) and the Review Committee on Genetic Manipulation-RCGM (BT/BS/17/371/2010-PID) were obtained for conducting the RT-PCR protocols on clinical patient samples, for the presence of SARS-CoV-2 RNA. Separate RCGM approval was obtained for testing the Toehold sensors (Document No BT/IBKP/035/2019).

## CODE AVAILABILITY

Custom software is deposited at Github (https://github.com/ShyamsundarR/silicasense) and available upon reasonable request.

## SUPPLEMENTARY FIGURE LEGENDS

Fig S1. A) IVTT assay performed on 19 shortlisted sensors was monitored using ONPG, a chromogenic substrate for lacZ. Fold change in Absorbance (420nm) is plotted for each sensor. Fold change was calculated with reference to the OFF state (sensor alone, no RNA added. B) IVTT assays performed on 4 selected sensors (1, 10, 12, 17) were monitored using CPRG, a red-shifted, chromogenic substrate for lacZ. Fold change in absorbance (576nm) is plotted for each sensor, with the addition of 10^13^ copies of Trigger RNA. Fold change is calculated relative to OFF state of sensor (sensor alone, no RNA added).

Fig S2. *Screen to identify the best NASBA primer pairs*. NASBA coupled with IVTT assay was performed using a fragment of SARS-CoV-2 RNA as template. A) 28 primer pairs were initially screened and tested for NASBA efficiency (primer sequences in Table S3). Template fragment contains the Trigger for sensor 12. Sensor 12 fused to lacZ was used for the IVTT assay. Data shows absorbance (576nm) over time for 10^8^ copies of template RNA. Response to 10^13^ copies of Trigger RNA (red) was used as a reference. Names of primer pairs are indicated in the index. Two types of forward primers were screened. 14 of the forward primers had a ‘short’ T7 sequence (TAATACGACTCACTATAGG) appended to them, while the other 14 forward primers had a longer version of T7 sequence (AATTCTAATACGACTCACTATAGGGAGAAGG) appended. The best primers pairs from here were shortlisted for phase 2 of testing. B) Phase 2 NASBA primer screen data is shown. Absorbance (576nm) over time for 10^4^ copies of template RNA is plotted. Response to 10^13^ copies of Trigger RNA (red) was used as a reference. Shortlisted primer pairs are indicated in the index (details of sequences are in Table S3).

Fig S3. *Sensitivity and specificity of assay for a range of human patient samples, with color read out*. NASBA coupled with IVTT assay performed on RNA extracted from nasopharyngeal swab samples. Samples had already been tested for the presence of SARS-CoV-2 RNA using standard real-time RT-PCR test and designated negative or positive (with accompanying Ct values shown here). Sensor 12 fused to lacZ was used for IVTT. Shown are Absorbance (576 nm) values over time. Samples are color coded based on their designation and Ct values. Response from patient samples with Ct values 16 to 25.4 are shown in panel A and response from patient samples with Ct values 26 to 35.7 are shown in Panel B. Ct values 36-38 (red) and -ve patients with Ct values > 45 (black) are shown. Caption indicates Ct values corresponding to individual curves. Sensor response can be used to distinguish COVID-19 positive from negative samples.

Fig S4. *Sensitivity and specificity of assay for a range of human patient samples with luminescence readout*. NASBA coupled with IVTT assay performed on RNA extracted from nasopharyngeal swab samples. Samples had already been tested for the presence of SARS-CoV-2 RNA using standard real-time RT-PCR test and designated negative or positive (with accompanying Ct values shown here). Sensor 12 fused to nano-lantern was used for IVTT. Shown are luminescence intensities values over time for patient samples with Ct values 16 to 25.4 (A) and samples with Ct values 28.7 to 35.7 (B). Response from patient samples Ct values 36-38 (red) and -ve patients with Ct values > 45 (black) are also shown for reference. Caption indicates Ct values corresponding to individual curves. Also shown are luminescence intensity values plotted against Ct values for patient samples at 30 minutes (C) and at 50 minutes (D), after the start of the IVTT assay post-amplification. Sensor response can be used to distinguish COVID-19 positive from negative samples.

Fig S5. *Time course analysis of NASBA amplification*. NASBA coupled with IVTT assay performed using a commercially available synthetic SARS-CoV-2 viral RNA control (Twist Biosciences) as template. NASBA amplification times ranged from 20, 40, 60 to 120 minutes. Sensor 12 fused to lacZ (color assay) or nano-lantern (luminescence assay) was used for IVTT. Representative data from 2 independent experiments (n=2) are shown. A) Image from a cell phone camera shows discernable color build up even with 100 copies of viral RNA as template and NASBA time of 60 minutes. B) Absorbance (576nm) over time shown for varying amounts of viral RNA (100 or 10^4^ copies) and varying NASBA times. C) Relative luminescence intensity over time for varying amounts of viral RNA (100 or 10^4^ copies of RNA). Trigger RNA at 10^13^ copies (red) is used as a positive control. Significant fold change in luminescence intensity is seen even at 100 copies of RNA template and 60 minutes of NASBA amplification.

## Notes

### Competing Interest Statement

The authors have declared no competing interest.

### Funding Statement

We are profoundly grateful to support from the Azim Premji Philanthropic initiative (APPI), PNB Housing CSR funds and IQVIA. We acknowledge support from the Department of Atomic Energy, Government of India and the National Centre for Biological Sciences-TIFR, under project no. 12-R&D-TFR-5.04-0800. Research in the A.R. lab is also supported by the Human Frontier Science Program research grant RGY0077/2019 and DBT/Wellcome Trust-India Alliance (IA/I/14/2/501521). A.G. thanks University of Hyderabad, the School of Life Sciences and Department of Biochemistry (UoH) for support. Research in AG lab is also supported by Scientific and Engineering Research Board (SERB), DST, Govt. of India (EMR/2017/005093).

### Summary of Updates

Additional results have been added to this updated manuscript.

