## Supplementary figures and images for "Engineered RNA biosensors enable ultrasensitive SARS-CoV-2 detection in a simple color and luminescence assay"

### Supplementary Figures

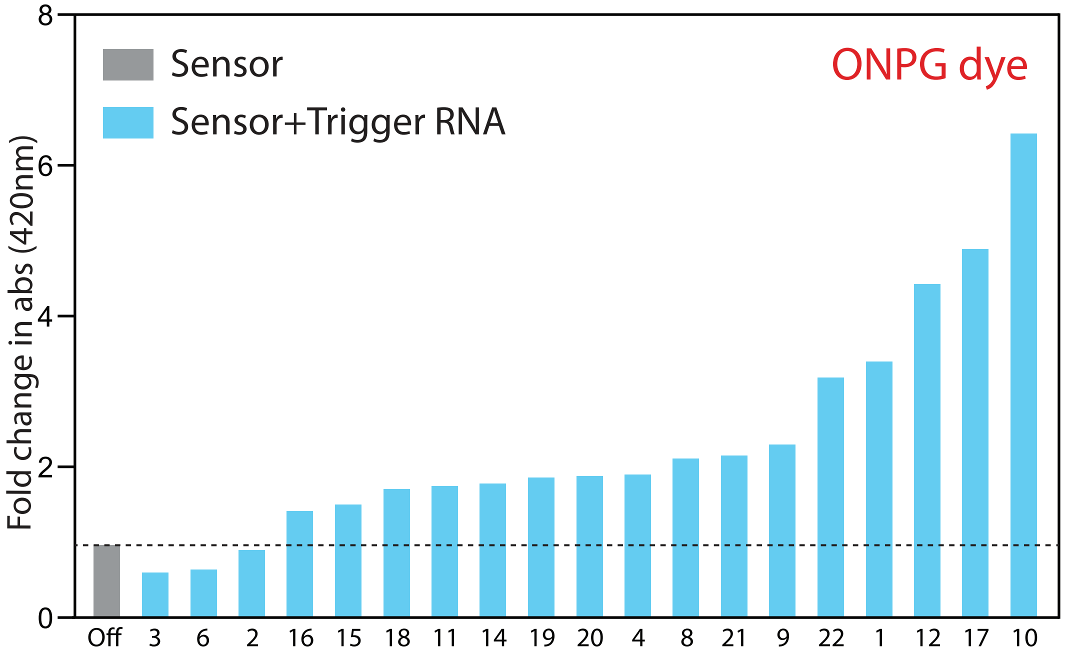

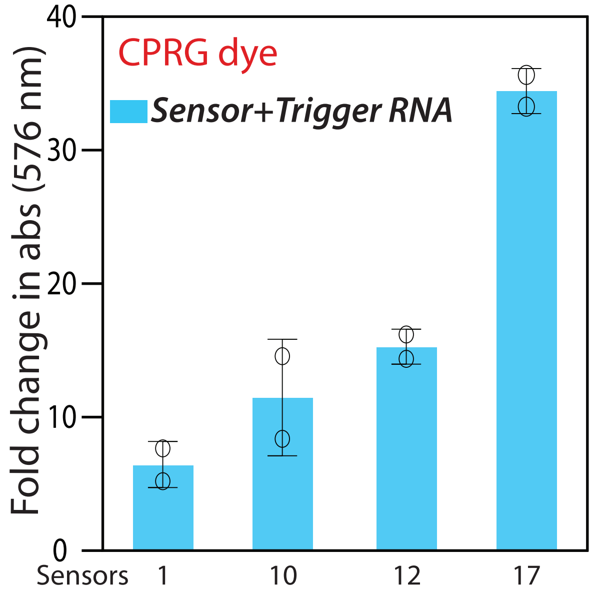

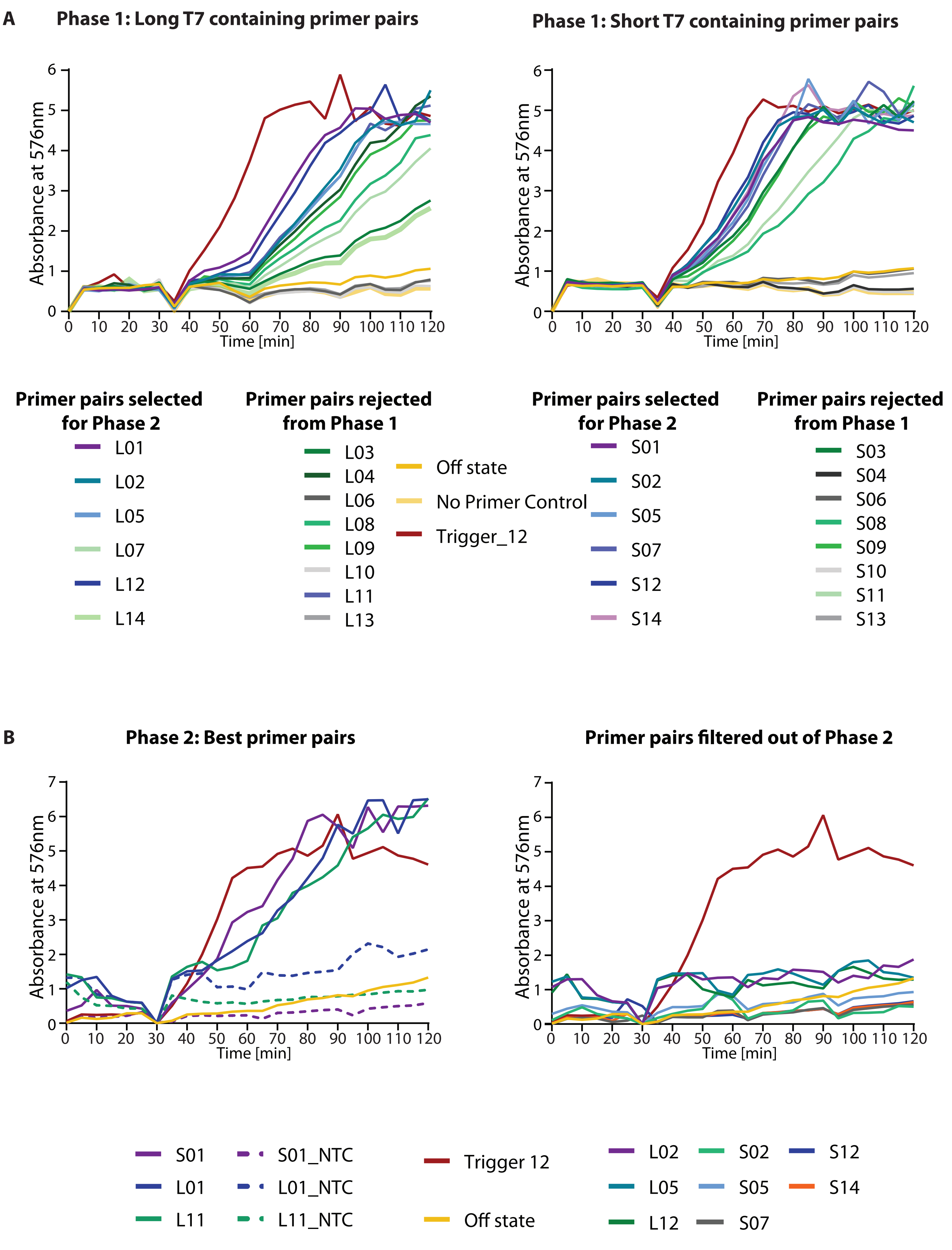

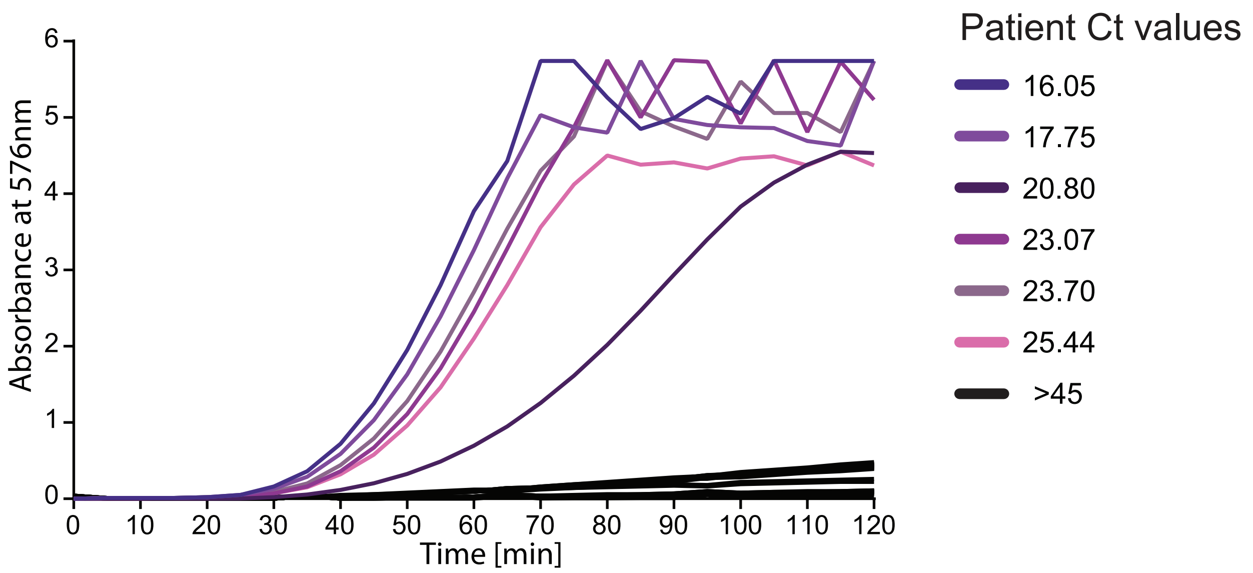

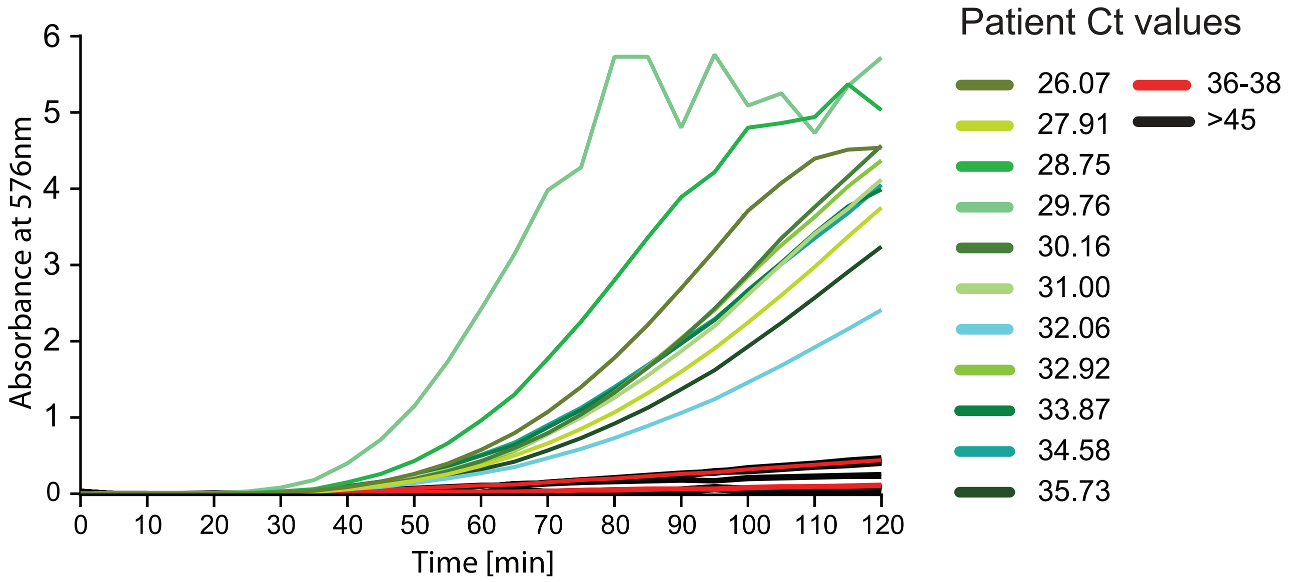

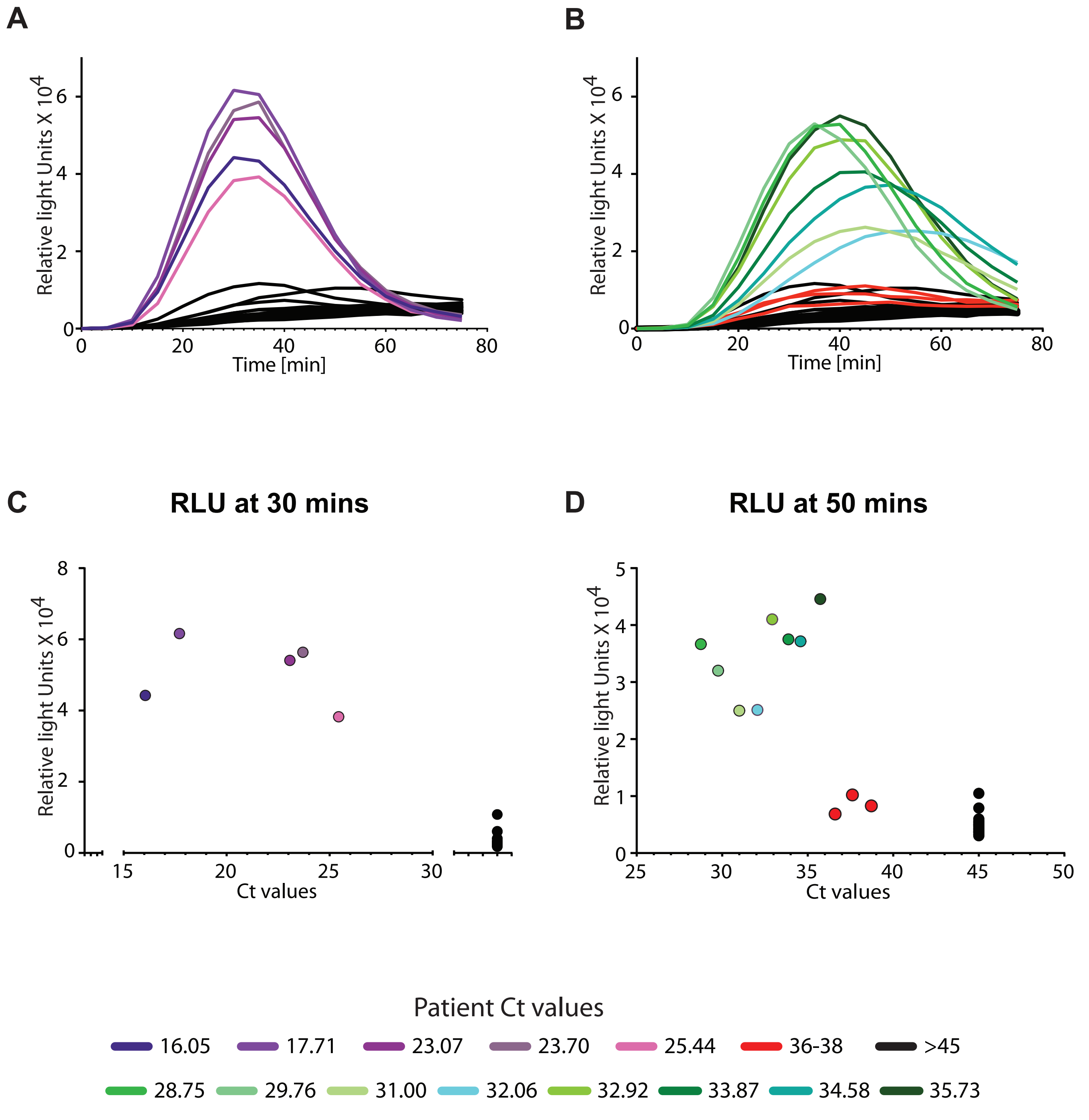

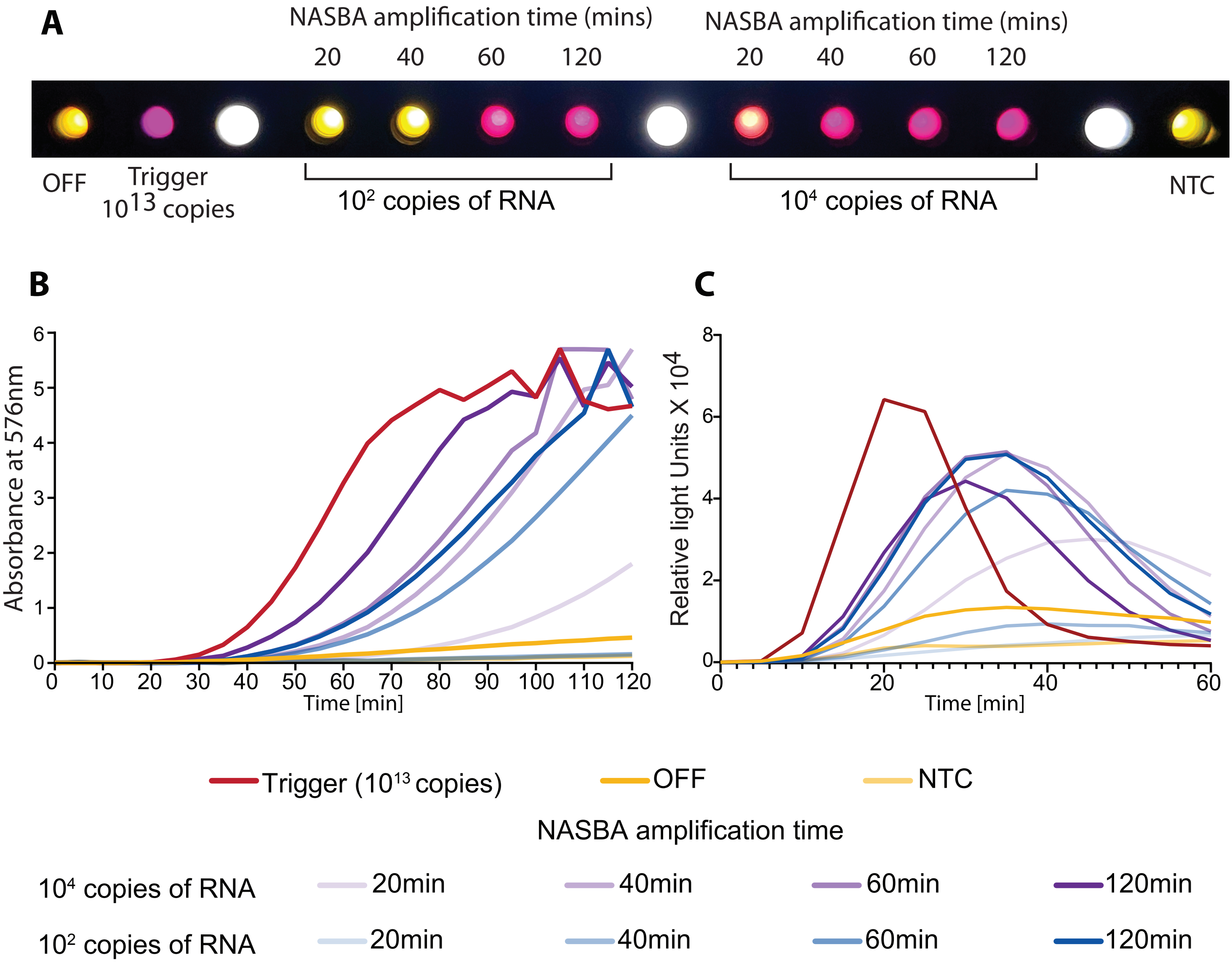
